# Evaluation of the secondary transmission pattern and epidemic prediction of COVID-19 in the four metropolitan areas of China

**DOI:** 10.1101/2020.03.06.20032177

**Authors:** Longxiang Su, Na Hong, Xiang Zhou, Jie He, Yingying Ma, Huizhen Jiang, Lin Han, Fengxiang Chang, Guangliang Shan, Weiguo Zhu, Yun Long

**Author notes:** **Corresponding authors** Guangliang Shan, Weiguo Zhu, and Yun Long.

## Abstract

Understanding the transmission dynamics of COVID-19 is crucial for evaluating its spread pattern, especially in metropolitan areas of China, as its spread can lead to secondary outbreaks outside Wuhan, the center of the new coronavirus disease outbreak. In addition, the experiences gained and lessons learned from China have the potential to provide evidence to support other metropolitan areas and large cities outside China with emerging cases. We used data reported from January 24, 2020, to February 23, 2020, to fit a model of infection, estimate the likely number of infections in four high-risk metropolitan areas based on the number of cases reported, and increase the understanding of the COVID-19 spread pattern. Considering the effect of the official quarantine regulations and travel restrictions for China, which began January 23∼24, 2020, we used the daily travel intensity index from the Baidu Maps app to roughly simulate the level of restrictions and estimate the proportion of the quarantined population. A group of SEIR model statistical parameters were estimated using Markov chain Monte Carlo (MCMC) methods and fitting on the basis of reported data. As a result, we estimated that the basic reproductive number, R_0_, was 2.91 in Beijing, 2.78 in Shanghai, 2.02 in Guangzhou, and 1.75 in Shenzhen based on the data from January 24, 2020, to February 23, 2020. In addition, we inferred the prediction results and compared the results of different levels of parameters. For example, in Beijing, the predicted peak number of cases was approximately 466 with a peak time of February 29, 2020; however, if the city were to implement different levels (strict, mild, or weak) of travel restrictions or regulation measures, the estimation results showed that the transmission dynamics would change and that the peak number of cases would differ by between 56% and ∼159%. We concluded that public health interventions would reduce the risk of the spread of COVID-19 and that more rigorous control and prevention measures would effectively contain its further spread but that the risk will increase when businesses and social activities return to normal before the end of the epidemic. Besides, the experiences gained and lessons learned from China are potential to provide evidences supporting for other metropolitan areas and big cities with emerging cases outside China.

## Introduction

The World Health Organization (WHO) named the virus 2019 novel coronavirus disease (COVID-19) and the novel virus severe acute respiratory syndrome coronavirus 2 (SARS-COV-2), which has attracted worldwide attention. The new coronavirus is a strain that has never been found in humans before. This virus can cause an acute respiratory disease, and common signs of infection include respiratory symptoms, fever, cough, shortness of breath, and dyspnea. In more severe cases, infection can cause pneumonia, severe acute respiratory syndrome, kidney failure, and even death(2020b).

According to WHO situation reports, the outbreak of COVID-19 has led to 79407 confirmed cases worldwide and 2622 deaths in 32 countries as of February 24, 2020, of which 64287 were from Hubei, China. Numerous cases have been reported in other areas outside Hubei, including metropolitan areas of Beijing (n=399) and Shanghai (n=335) inside China, as well as other countries outside China, such as South Korea (n=833), Japan (n=144), and Italy (n=124). With the continuously increasing number of cases, understanding the spread pattern of COVID-19 and monitoring spikes in the number of cases are crucial steps to provide evidence that can guide public health intervention strategies and healthcare policy making.

Several mathematical models and data analysis approaches attempting to estimate the transmission of COVID-19 have been recently reported(Hermanowicz, 2020;Imai et al., 2020;Liu et al., 2020a). Public health interventions and transportation restriction effects for disease transmission have also been evaluated in some studies(Tang et al., 2020;Zhao et al., 2020). Some studies indicated that public intervention measures greatly mitigate the final size of the epidemic, and shift the turning point about 24 days before the turning point without these measures(Liu et al., 2020b). Some noted that travel restrictions would not affect much unless combined with a 50% or higher reduction of transmission in the community(Chinazzi et al., 2020). And a report from Imperial College COVID-19 Response Team concluded that the intensive intervention or something equivalently effective, such as combining home isolation of suspect cases, home quarantine of those living in the same household as suspect cases, and social distancing of the elderly and others at most risk of severe disease, could reduce transmission; however, it will need to be maintained until a vaccine becomes available, and the team also predicted that transmission will quickly rebound if interventions are relaxed so it requires multiple interventions to be combined to have a substantial impact on transmission(Ferguson et al., 2020). Besides, to predict the outbreak size and time, researchers have published many different results for forecasting when the outbreak will peak in different areas(Read et al., 2020;Wu et al., 2020). These models are certainly useful to understand the emerging trends of COVID-19. However, there are several challenges to such timely analyses and forecasting. Due to barriers, such as the disease incubation period, asymptomatic infection, diagnosis testing capacity, overloaded medical staff and complicated reporting processes, there can be a delay or missed reporting in this evolving situation regarding the confirmation of cases. Furthermore, the adopted models have mostly been complicated with many pre-settings or assumptions or parameter values that are likely not accurate. Although some modeling approaches can estimate parameter values through statistical methods, they can only contribute a rough simulation for the modeling. As a result, those studies achieved different prediction results by using different methods and datasets.

To achieve a relatively objective judgment, given that that this new disease and complicated situation has many unknown factors, we used mathematical modeling methods to characterize COVID-19 transmission and used multiple datasets for ensuring the data reliability. Since individual data sources may be biased or incomplete, according to related studies, the use of multiple data sources rather than a single dataset can enable a more robust estimation of the underlying dynamics of transmission(Funk and Eggo). Therefore, we investigated and collected data from four sources, including released data and official daily reports from commercial technology companies, academic institutes, authorities or local healthcare commissions, and the World Health Organization, to minimize the resulting errors caused by potentially biased single data sources. The data were obtained from the Beijing Municipal Health Commission (BMHC)(Decker, 2010), Shanghai Municipal Health and Family Planning Commission (SMHFPC)(2020e), Health Commission of Guangdong Province(2020c), National Bureau of Statistics of China (NBSC)(2020d), Baidu Migration Big Data Platform (BMBDP)(2020a), Center for Systems Science and Engineering (CSSE) of Johns Hopkins University(Dong et al., 2020), and WHO coronavirus disease (COVID-2019) situation reports(Organization, 2020c). Considering that the cases detected in these four cities were all imported and secondary transmission cases, and based on the reported data available after January 20, 2020, Chinese authorities have implemented prevention measures in these cities to contain the outbreak and prevent the disease from spreading; thus, we considered the secondary transmission pattern of COVID-2019 to be different than the early spread pattern in Wuhan, where the virus was rampantly transmitted without any prevention measures. Therefore, we collected data from January 24, 2020 (Chinese New Year’s Eve) to February 23, 2020 to give an overall objective estimation of COVID-19 development in 4 high-risk metropolitan areas of China: Beijing, Shanghai, Guangzhou, and Shenzhen. We estimated how COVID-19 human-to-human transmission occurred in these large cities, which have developed considerable cases. We further used these estimates to forecast the potential risks and development trends of these four metropolitan areas inside China.

## Methods

To evaluate the COVID-19 spread pattern and estimate its transmission in 4 metropolitan areas, we used an adjusted SEIR model with data. We only considered human-to-human transmission in our models.

### Adjusted SEIR model for COVID-19

The SEIR model is a deterministic metapopulation transmission model in which the population is divided into four classes: S (susceptible, people who are likely to be infected), E (exposed, people who are exposed), I (infectious, people who are infected) and R (removed, recovered and dead persons). We assumed that the epidemic risk started with infectious cases on February 3, 2020, when authorities announced that people were returning to work after the Chinese Spring Festival holiday. Therefore, we modeled a period beginning on February 3, 2020. The SEIR model state transition is shown in Figure 5. In our estimation, the entire population was initially susceptible since COVID-19 is an emerging new infectious disease and not all people have immunity against it. In January (before Chinese New Year),there were estimated 3.246, 2.847, 3.430, 3.271 million people flow out from Beijing, Shanghai, Guangzhou and Shenzhen, respectively. We took this outflow number out from these four cities initial population and assume they inflow back after Chinese New Year by February 17, 2020. We estimated the initial exposed population using number of confirmed cases during the next 7 days. We assumed that the median incubation period was 5-6 days (ranging from 0-14 days) based on the WHO report(Organization, 2020a).

Based on the basic SEIR model, we further considered the influence of multiple factors on the transmission pattern as the situation unfolded, including public health intervention measures, people’s self-protection behaviors, the diagnosis rate, population flow, etc.

Assuming that public health interventions contributed to the control of the dynamics of the epidemic, we incorporated an parameter that indicates the changes in the population flow into the model. According to the inflow index, outflow index and urban daily adjusted index of the travel intensity from the Baidu Migration Big Data Platform, for the period from January 24, 2020 to February 23, 2020, we inferred that the people’s activity was obviously lower than the normal level for the same period last year. Furthermore, considering the Spring Festival population flow and those returning to work after the holiday (officially announced as February 3, 2020), we regarded that the risk for these 4 metropolitan areas grows with the inflow population increase starting on February 3, 2020, an average introduced number of cases were counted into the model.

We also estimated the parameter values within these cities using the MCMC method. Cases in the reported data and other sources reported between January 24, 2020 and February 23, 2020 were used to adjust the model. Considering the possible complex influencing factors, we proposed an adjusted SEIR model for COVID-19 estimation, as displayed in Figure 6.

In the adjusted SEIR model, we considered the inflow of the city’s population, so the total number of people was not fixed, and the population was divided into seven classes: S (susceptible, people who are likely to be infected), E (exposed, people who are exposed), I (infectious, people who are infected), R (recovered and dead persons), Sq (quarantined susceptible persons), Eq (isolated exposed persons) and Iq (isolated infected persons). The transmission dynamics are governed by the following equations:

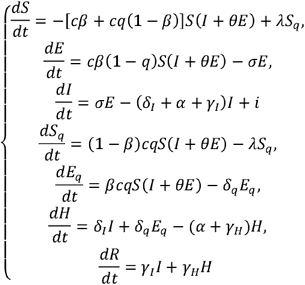

where q is the quarantined proportion of exposed individuals, β is the transmission probability per contact and c is the contact rate which defines how many people are contacted with an infected person per day, i is the estimated infected people within the inflow population each day. The quarantined infected people moved to the compartment E_q_ at a rate of βcq, with the quarantined uninfected people moved to the compartment S_q_ at a rate of (1–β)cq. Those who were not quarantined, if infected, moved to compartment E at a rate of βc(1-q). θ is the transmission capability between the latent and the infected population. According to the reported results of related work, the transmission capability of the people in the incubation period and the diagnosed infected patients are similar(Qiu, 2020), we assume that. is θ= 1 λ is the the transition rate from the quarantined to susceptible, σ is the transition rate from the exposed to the infected population, α is the mortality rate, δ_I_ is the transition rate from the infected population to quarantined infected population, and γ_I_ recovery rate of the infected population. δ_q_ is the transition rate from the quarantined exposed population to the quarantined infected population, and γ_H_ is the recovery rate of the quarantined infected population.

### Parameter estimate methods

The MCMC method is a commonly used algorithm in modern statistical calculations. This algorithm provides an effective tool for establishing statistical models and is widely used in Bayesian calculations of complex statistical models(Endo et al., 2019). We used the MCMC method and Metropolis-Hastings(MH) algorithm sampling(Hastings, 1970), with a normal distribution as the recommended distribution, estimated the parameters of the modified SEIR model, to obtain the baseline estimation of parameters, and incorporated the data collected from infectious disease reports into the above statistical inference, and simulated the process of infectious disease transmission to further fix some parameters on the basis of fitting reported data. Using Beijing as an example, the parameter estimates and initial values of the SEIR model are listed in Table 3.

**Table 1.** The effects of the contact rate on the peak time and peak value with an estimated value.

| Areas | Parameter c | 2c | 1.5c | c | 0.8c | 0.6c |
| --- | --- | --- | --- | --- | --- | --- |
| Beijing | Days to Peak | 16 | 19 | 27 | 31 | 41 |
|  | Peak Value | 642 | 608 | 476 | 399 | 286 |
| Shanghai | Days to Peak | 16 | 19 | 25 | 29 | 40 |
|  | Peak Value | 592 | 545 | 473 | 406 | 309 |
| Guangzhou | Days to Peak | 16 | 18 | 25 | 28 | 40 |
|  | Peak Value | 515 | 481 | 403 | 353 | 279 |
| Shenzhen | Days to Peak | 17 | 20 | 25 | 32 | 45 |
|  | Peak Value | 688 | 542 | 487 | 478 | 377 |

**Table 2.** The effects of the quarantined rate of exposed individuals on the peak time and peak value.

| Areas | Parameter $q$ | $2q$ | $1.5q$ | $q$ | $0.8q$ | $0.6q$ |
| --- | --- | --- | --- | --- | --- | --- |
| Beijing | Days to Peak | 20 | 22 | 27 | 29 | 32 |
|  | Peak Value | 259 | 325 | 476 | 576 | 742 |
| Shanghai | Days to Peak | 20 | 22 | 25 | 27 | 29 |
|  | Peak Value | 290 | 378 | 473 | 662 | 886 |
| Guangzhou | Days to Peak | 20 | 23 | 25 | 27 | 29 |
|  | Peak Value | 389 | 352 | 403 | 643 | 842 |
| Shenzhen | Days to Peak | 18 | 21 | 25 | 25 | 27 |
|  | Peak Value | 272 | 329 | 487 | 598 | 789 |

**Table 3.**
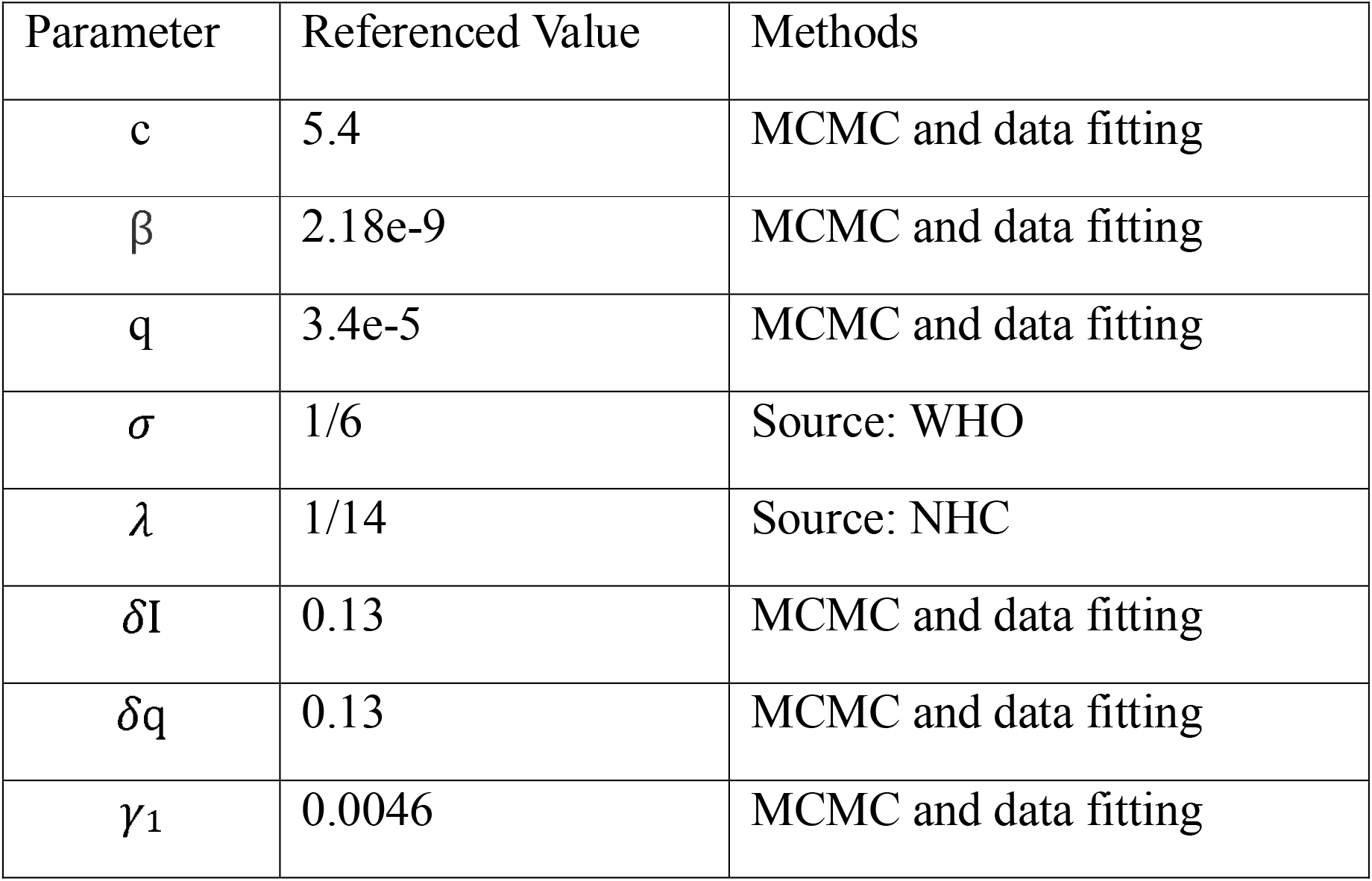
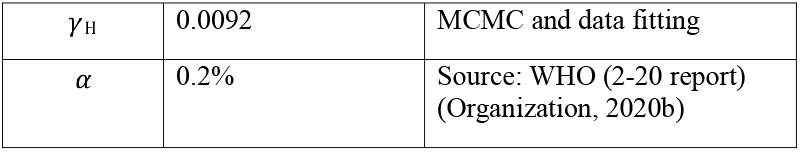
Parameters and initial values for the adjusted SEIR model (Beijing)

In addition, to simulate the contact rate for model estimation, we used urban travel index data from Baidu, a major internet company in China that hosts the popular navigator app Baidu Maps, which indirectly monitors the real-time urban travel intensity and population flow. The Baidu index of travel intensity and population flow was converted into the corresponding coefficients for the contact rate and the quarantined susceptible population. In terms of Baidu index, we simulated people’s activity level by comparing our observed period (under strict interventions) with normal level in same period last year, we also consider the assumption scenario that when people returning back to work (limited interventions), accordingly, we added the coefficients (0.6c, 0.8c, c, 1.5c, 2c) for the baseline contact rate to compare different effectiveness of interventions. Similarly, the coefficients were added to baseline quarantine proportion (0.6q, 0.8q, q, 1.5q, 2q).

### Basic reproduction number R_0_ estimates

At the onset, when all people are susceptible, R_0_ is defined as the average number of new infections directly caused by a case in a population of people who are all susceptible. Given the model structure includes quarantine and isolation, we used the next generation matrix to derive a formula for the basic reproduction number after public health interventions were executed, the principal eigenvalue of the next generation matrix is the expectation of population growth and the equation is as follows and the parameter definition is same with adjusted SEIR model.

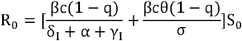

## Results

### Data Characterization

To characterize the overall epidemic size and dynamics, Figure 1 shows the epidemic curve of COVID-19 cases identified in Beijing, Shanghai, Guangzhou and Shenzhen from January 24, 2020 to February 23, 2020.

**Figure 1.**
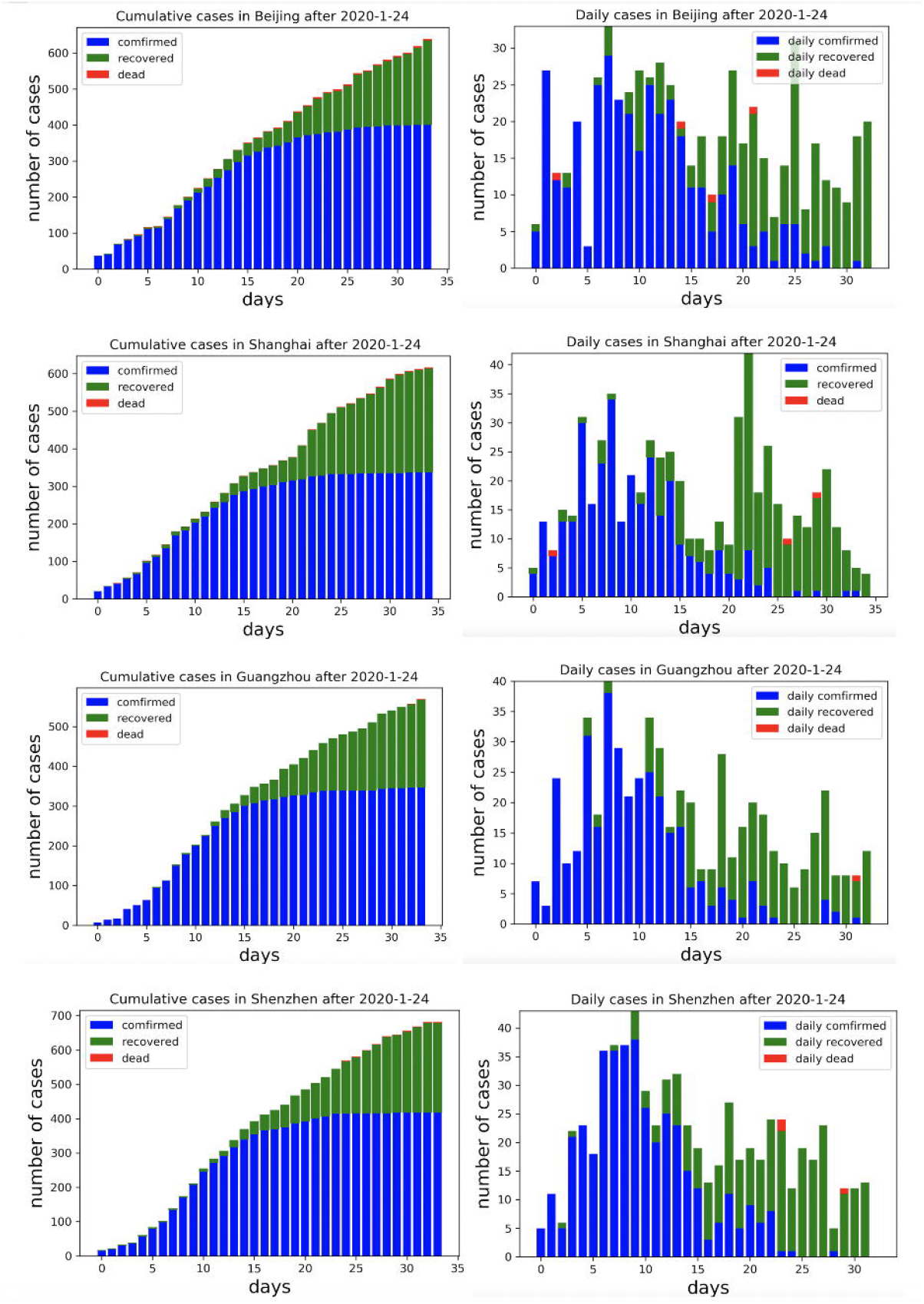
Cumulative and daily reported cases in 4 metropolitan areas in China.

### Adjusted SEIR Model Estimation

We summarized and interpreted the transmission dynamics of COVID-19 in the 4 municipal areas. The adjusted SEIR model was used to predict cases in Beijing, Shanghai, Guangzhou and Shenzhen, and Figure 2 shows the comparisons between the predicted results and actual results. The results are based on an assumption of no further imported cases to these cities since China implemented strong regulation measures during the observation period.

**Figure 2.**
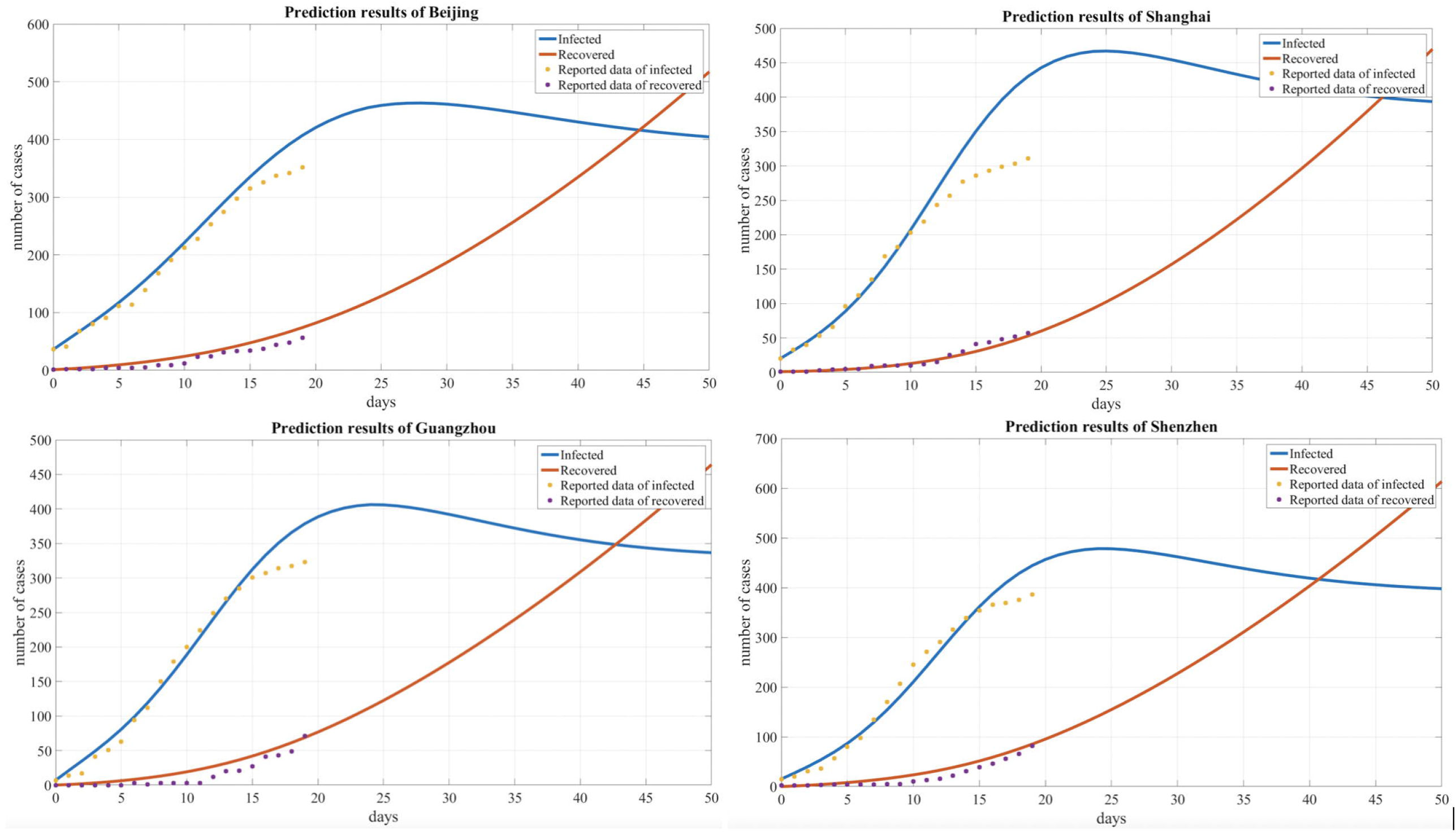
Comparison of the predicted and actual numbers of infected and cured people in 4 cities.

Based on our observations from the data shown in Table 1 and Figure 3 below, we also found that the number of infected individuals changed with different levels of public health interventions and that strict interventions could decrease the peak number of infected individuals compared with the scenario of weak interventions; accordingly, we used different contact rates to reflect the different levels of interventions. The baseline contact rate was derived by the MCMC method, and the results show that reducing the contact rate persistently decreased the peak value or could delay the peak. In addition, with strict public intervention, the number of infected individuals eventually decreased and the peak appeared sooner than it would with weak intervention methods. After February 3, 2020, as people returned to work after a holiday, many people returned to these cities, which was inferred from the Baidu transportation index. We added this information into the risk factors for the contact rate (1.5c, 2c). Accordingly, the number of infected individuals increased compared with the scenario of a decreased contact rate (0.8c, 0.6c).

**Figure 3.**
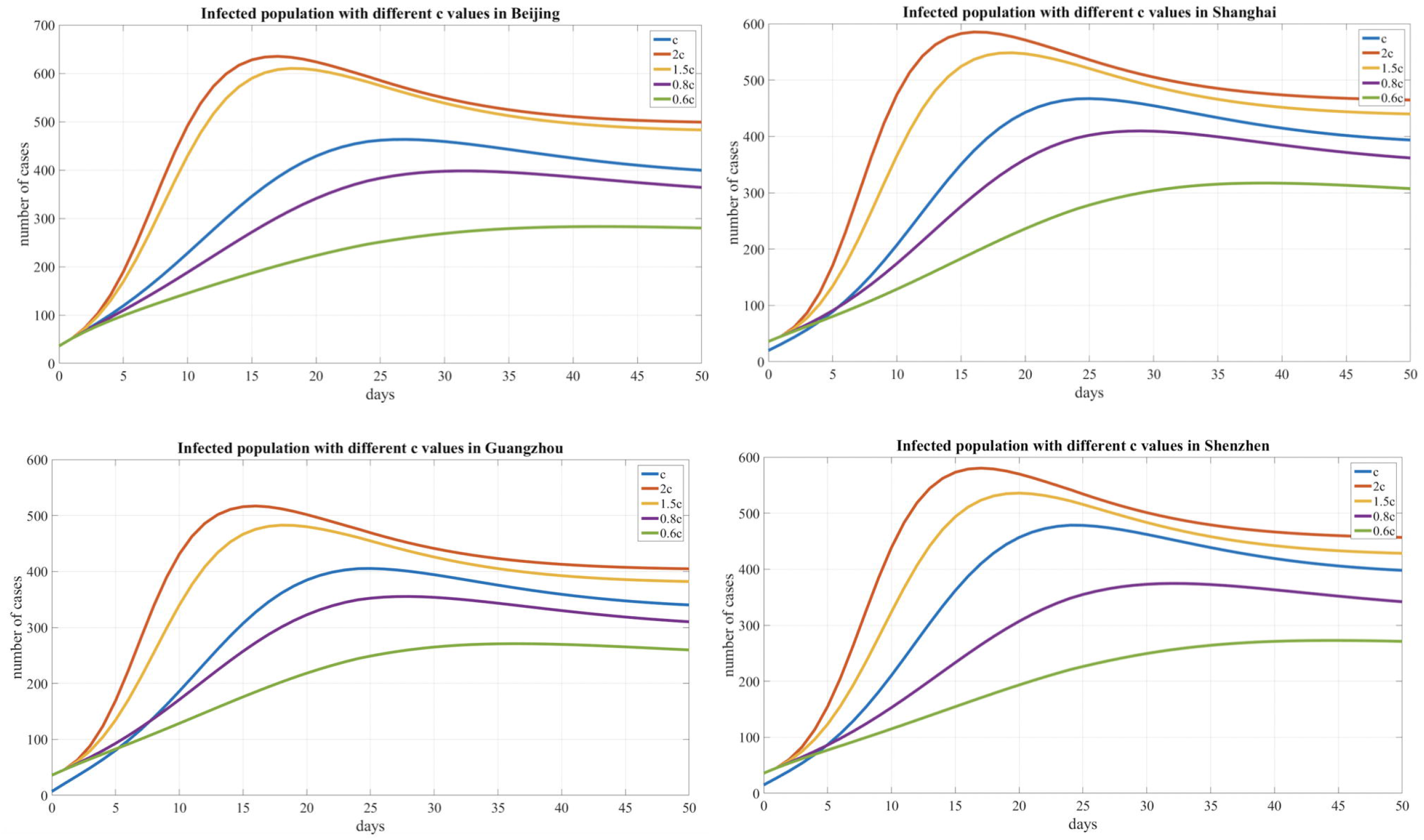
Infected population curves with different contact rates for 4 cities.

**Figure 4.**
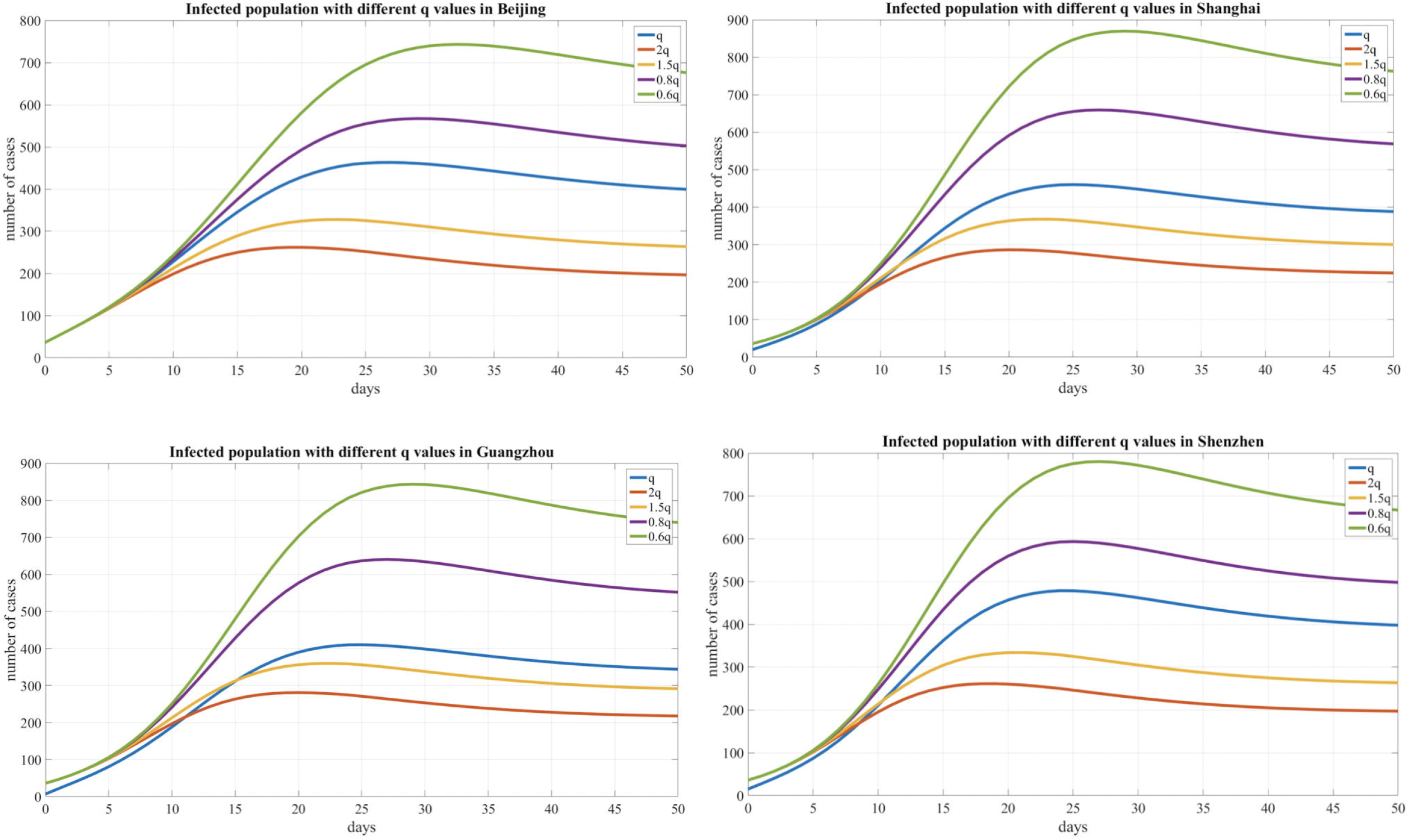
Infected population curve with different quarantined proportion of exposed individuals in 4 cities.

**Figure 5.**
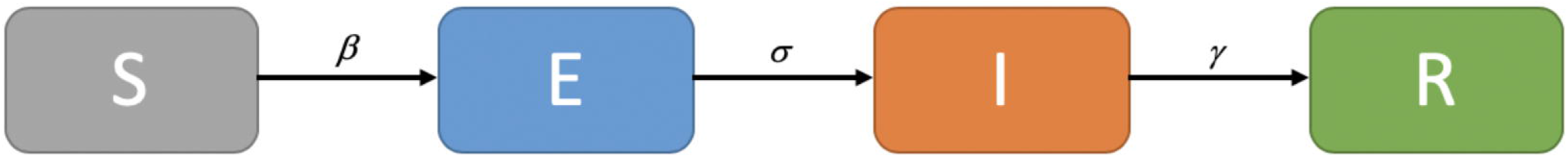
SEIR Model

**Figure 6.**
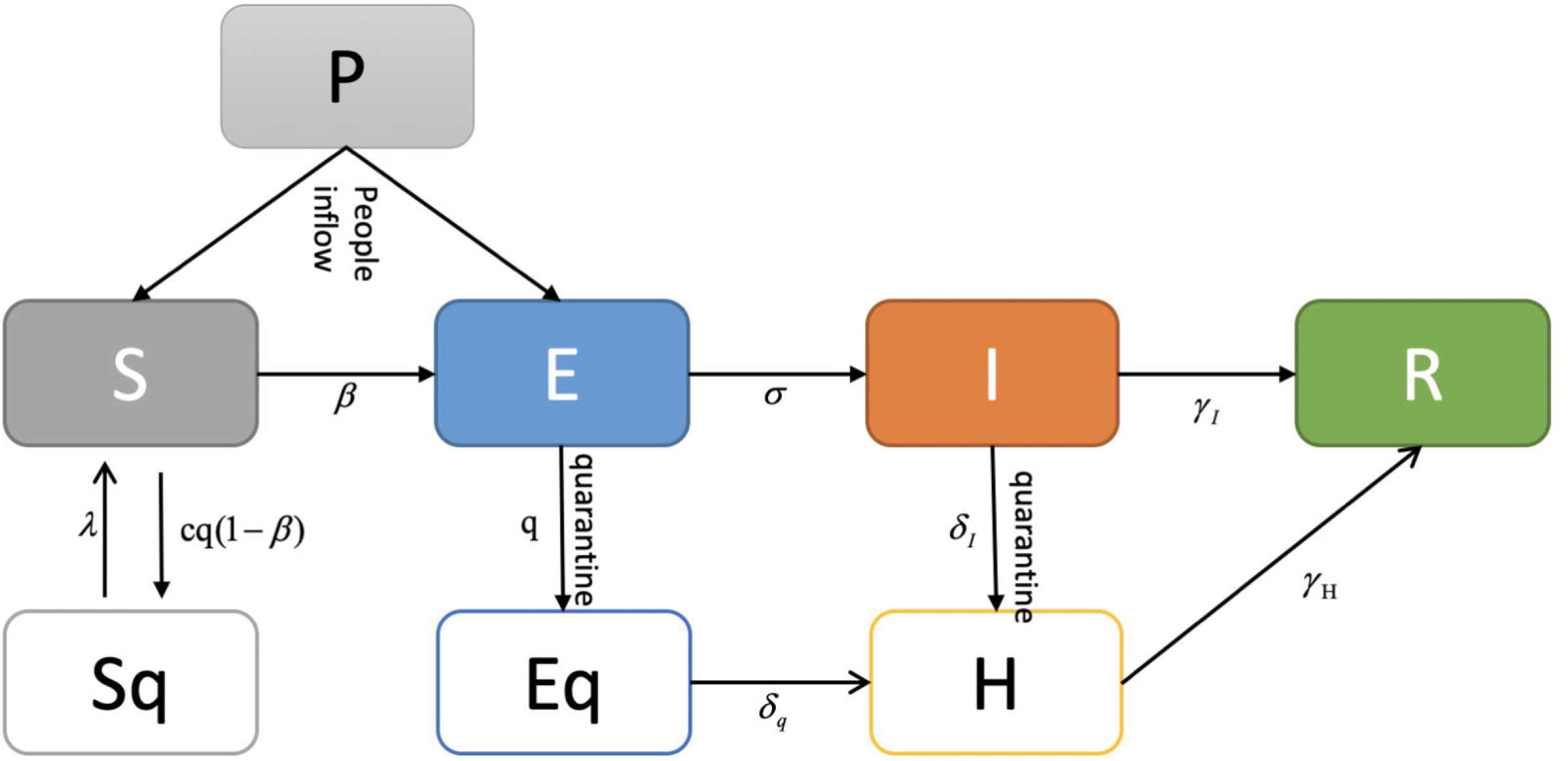
Adjusted SEIR model for COVID-19

In addition, we compared the transmission dynamics with different quarantined proportion of exposed individuals, which reflects the contact tracing capability and management efforts of local governments, and the results show that reducing the quarantined proportion of exposed individuals (0.8q, 0.6q) led to an increase in the peak value and delayed the peak time. Conversely, the peak value decreased and an earlier peak time occurred with a higher quarantined proportion of exposed individuals (2q, 1.5q).

### R_0_ Estimation Results

We used the MCMC method to fit the model and adopted an adaptive MH algorithm to carry out the MCMC procedure. As a result, we inferred R_0_= 2.91, 2.78, 2.02, and 1.75 for Beijing, Shanghai, Guangzhou and Shenzhen, respectively.

## Discussions

Our analysis results strongly demonstrate that reducing secondary infections among close contacts would effectively limit human-to-human transmission, and public health measures, such as the rapid identification of cases, tracing and following up with people who had contact with an infected person, infection prevention and control in health care settings, and the implementation of health measures for travelers, can greatly prevented further spread of the disease.

The documented COVID-19 reproduction numbers range from 2.0 to 4.9(Shen et al., 2020;Wu et al., 2020;Zhao et al., 2020), which are based on cases that developed during different transmission phases and in different areas. For instance, the R_0_ in Wuhan was obviously higher than that in other cities outside Wuhan during the timeframe analyzed. Furthermore, after implementing the prevention measures employed by the Chinese government and local authorities, we regarded the inferred R_0_ results of the four cities as reasonable and interpretable.

In this study, we aimed to monitor COVID-19 trends after cases were imported into other cities and countries and estimate the spread pattern by mathematical modeling, which can be helpful for evaluating the potential risk and severity of new outbreaks. The results of our study show that, for four metropolitan areas of China, the containment measures were effective control at that time; however, it is imperative to raise awareness in the population and prevent potential outbreak risks any time. The study has limitations. The present reported data are insufficient to understand the full epidemiological pattern of COVID-19 transmission and new potential outbreaks. For example, the estimates in this manuscript have a certain extent of uncertainty and delays due to the limitations in reporting mechanisms over the course of the natural history of the cases, the impact of other potential asymptomatic cases and some unreported cases. Some studies conducted assumption that a small fraction, 20%, were not reported(Liu et al., 2020b) and others reported the estimated asymptomatic proportion was 17.9%(Mizumoto et al., 2020) or 60%(Qiu, 2020).Evidently, such asymptomatic infectious cases are not fully reported by authorities. However, some studies suggested crowdsourced data could be compiled and analyzed as an complementation of officially released data, this perhaps help improving the analysis results(Arbia, 2020;Leung and Leung, 2020;Sun et al., 2020). In the future, we will evaluate how will the number of unreported cases influence the severity of the epidemic and consider involving these unreported data to our mathematical model.

As concluded from the WHO-China Joint Mission report(Organization), the COVID-19 transmission dynamics are inherently contextual, as are the dynamics for any outbreak, and people worldwide need to work together to defend against this disease, including 1) to enhance the understanding of the evolving COVID-19 outbreak in China and the nature and the impact of ongoing containment measures; 2) to share knowledge on the COVID-19 response and preparedness measures being implemented in countries affected by or at risk of importations of COVID-19; 3) to generate recommendations for adjusting COVID-19 containment and response measures in China and internationally; and 4) to establish priorities for a collaborative program of work, research and development to address critical gaps in knowledge, responses, readiness tools and strategies.

As a consequence of our study, we concluded that the outbreak could be greatly reduced by strict public health interventions. The public intervention strategies and implemented protection measures conducted in these four areas may help provide epidemiological suggestions to governments that guide measures for the international cases that are rapidly emerging.

## Data Availability

The data used to support the findings of this study were provided by Dr. Longxiang Su and Dr. Na Hong under license and thus cannot be made freely available. Access to these data will be considered by the author upon request.

## Author Contributions

L. S and N. H and X. Z contributed equally. W. Z and Y. L and G. S take responsibility for the integrity of the work as a whole, from inception to published article; N. H and L. S were responsible for study design and conception; F. C and L. H collected and cleaned the data; J. H, Y. M and H. J were responsible for data modeling and analysis; W. Z, G. S and X. Z interpreted the results; L. S and N. H and X. Z. drafted the manuscript. All authors revised the manuscript for important intellectual content.

## Declarations

Author Na Hong, Jie He, Yingying Ma, Lin Han and Fengxiang Chang were employed by the company Digital China Health Technologies Co. Ltd., Beijing, China. The remaining authors declare that the research was conducted in the absence of any commercial or financial relationships that could be construed as a potential conflict of interest.

